# Hair concentrations of Δ-9-tetrahydrocannabinol (THC) and cannabidiol (CBD) in cannabis consumers psychiatric patients

**DOI:** 10.1101/2022.04.13.22273751

**Authors:** Yann Barguil, Laura Chiaradia, Guy Southwell, Jean-Yves Charlot

## Abstract

Among young consumers of cannabis, a brief psychotic disorder (BPD) can be either the clinical manifestation of acute cannabis psychosis (ACP) or an announcement of schizophrenia’s onset. Clinicians are faced with the difficulty of making a differential diagnosis between disorders of the schizophrenic field and disorders induced by cannabis. To date, no clinical or even less paraclinical criteria have made it possible to differentiate syndromes whose prognoses and management are different. Since 2010, we measured delta-9 tetrahydrocannabinol (THC) and cannabidiol (CBD) concentrations in head hair among New Caledonian patients, all cannabis consumers (n = 256). We wanted to determine if these patients, cannabis users, suffering from different mental pathologies, present particular phenotypes of capillary cannabinoid concentrations (THC and CBD). At the time of initial psychiatric consultation, a sample of 3 cm proximal length of head hair was prepared for analysis, and THC and CBD were then assayed by Gas Chromatography coupled with Mass Spectrometry (Limit Of Quantitation: 0.05 ng/mg). At the end of the 6 months medico-psychologic follow-up from the initial evaluation, four groups of cannabis users were identified according to the final psychiatric diagnosis: control, acute cannabis psychosis (ACP), chronic psychosis (CP), and other personality disorders (OPD) groups. In this study, a high hair level of THC detected (> 0.7 ng/mg) associated with a low hair CBD/THC ratio (< 0.26) are two parameters that taken together could be good markers of CP development. For OPD and ACP, hair CBD/THC ratios were higher in the ACP group (> 0.43) than in the OPD group (< 0.32). This study highlights, once again, the protective role of CBD against the deleterious effects of THC. In association with clinical evaluation, this toxicological approach could be helpful for psychiatric diagnosis and would allow early management of BPD in cannabis consumers. For a consumer who does not present with a psychiatric disorder, it could give an information about the possibility of belonging to a group of patients at high risk of psychiatric decompensation. This provides an additional argument for efforts to control cannabis consumption by patients.

## Introduction

The psychotoxicity of *Cannabis sativa* var. indica is well-established [1]. It can lead to the development of both short -and long-term psychiatric disorders or aggravate pre-existing psychiatric disorders [2-8]. Among young consumers of cannabis, a brief psychotic disorder (BPD) can be either the clinical manifestation of acute cannabis psychosis (ACP) or an announcement of schizophrenia’s onset [9, 10]; therefore, clinicians are faced with the difficulty of making a differential diagnosis between disorders of the schizophrenic field and disorders induced by cannabis [10]. The clinical presentation is of the same order, and only the evolution of disorders makes it possible to differentiate between these entities. If care is identical in the immediate future, it must be reconsidered in the medium-and long-term. To date, no clinical or even less paraclinical criteria have made it possible to differentiate syndromes whose prognoses and management are different [11]. Close collaboration between psychiatrists, medical biologists, and analysts has made it possible to propose a simple protocol that allows assistance with the therapeutic approach.

## Patients and method

To assess the extent of cannabis addiction and/or to monitor compliance with abstinence programs by consumers, since 2010, we measured delta-9 tetrahydrocannabinol (THC) and cannabidiol (CBD) concentrations in head hair among New Caledonian patients, all cannabis consumers (n = 256). This is a well-established clinical practice [12]. A lock of the posterior vertex region of the head hair was taken and cannabinoids of interest (THC and CBD) were measured by gas chromatography coupled with mass spectrometry (GC-MS) on the proximal 3 cm, representing approximately the last 3 months period [13]. Hair cannabinoids is a useful test to detect heavy cannabis use [12, 14, 15].

We decided to conduct a prospective study; indeed, some of these patients also showed clinical signs that may be related to different mental disorders. These patients then benefit from psychiatric follow-up, and psychiatric diagnosis is made according to the Diagnostic and Statistical Manual of Mental Disorders, Fifth Edition, Text Revision (DSM-5-TR) after 6 months of follow-up [16].

We wanted to determine if these patients, cannabis users, suffering from different mental pathologies, present particular phenotypes of capillary cannabinoid concentrations (THC and CBD). In other words, it would be possible to make a first diagnosis just after the initial psychiatric evaluation, thus making it possible to overcome a significant delay (3–6 months), delaying the most appropriate care. Appropriate care and early therapeutic management are important if the pathology responsible for mental disorders is chronic psychosis. Schizophrenia, for example, is a progressive disease, and a delay in therapeutic management compromises the chances of socio-professional reintegration [11].

The 256 participants included in this study were over the age of 16 years, consumers of cannabis, and tested positive for cannabis in urine. The included patients had previously received oral and written information about the objective of the study, and consent was obtained. People under the age of 16 years, people with bleached hair, and people with mental disabilities were excluded from the study.

At the time of initial psychiatric consultation, a sample of 3 cm proximal length of head hair was prepared for analysis, and THC and CBD were then assayed by Gas Chromatography coupled with Mass Spectrometry (GC-MS) (LOQ: 0.05 ng/mg) [13].

At the end of the 6 months medico-psychologic follow-up from the initial evaluation, four groups of cannabis users were identified according to the final psychiatric diagnosis.

Group 1: 62 patients hospitalized for medical or surgical reasons (Control).

Group 2: 32 patients undergoing psychiatric follow-up for of acute cannabis psychosis (ACP).

Group 3: 92 patients undergoing psychiatric follow-up for chronic psychosis (CP).

Group 4: 70 patients with other personality disorders (OPD) (borderline personality disorder, mood disorder, etc.).

### Statistical analyses of data

Capillary CBD and THC concentrations were statistically analyzed. GraphPad Prism 8 software was used for statistical analyses. Samples for which CBD and/or THC were not detected were considered in this study because these patients are known to be cannabis consumers (urine samples were positive for THC-COOH prior to hair sampling). A threshold value was arbitrarily set at 0.0024 ng/mg for the calculation of ratios because we verified cannabis positivity in urine samples. Molecules of interest were present in trace amounts but were not quantifiable by the automation used in this study. The CBD/THC ratios were calculated for each patient.

Data were described for each group of patients to obtain the mean and a confidence interval of 95 %. These data were used to generate a decision tree, which may help guide clinicians in the early diagnosis of chronic psychosis in cannabis-consuming patients presenting with a brief psychotic disorder (BPD).

The data obtained in each group regarding the concentrations of capillary THC, CBD, and CBD/THC ratios were compared. Comparisons were made using Tukey’s multiple comparisons test (α = 0.05), and all groups were compared to each other for each variable.

## Results

In some patients, THC and/or CBD were not detected in their hair. Nevertheless, all data obtained were used for this study, as all patients are known to be cannabis consumers. To exploit these data, undetected CBD or THC were arbitrarily fixed at 0.0024 ng/mg. Indeed, the limit of quantification (LOQ) for automation was 0.05 ng/mg. Detected but not quantifiable CBD or THC were 0.025 ng/mg (half of LOQ). In the histogram below (*figure 1*), represents the data obtained for each group in this study.

**Figure 1.**
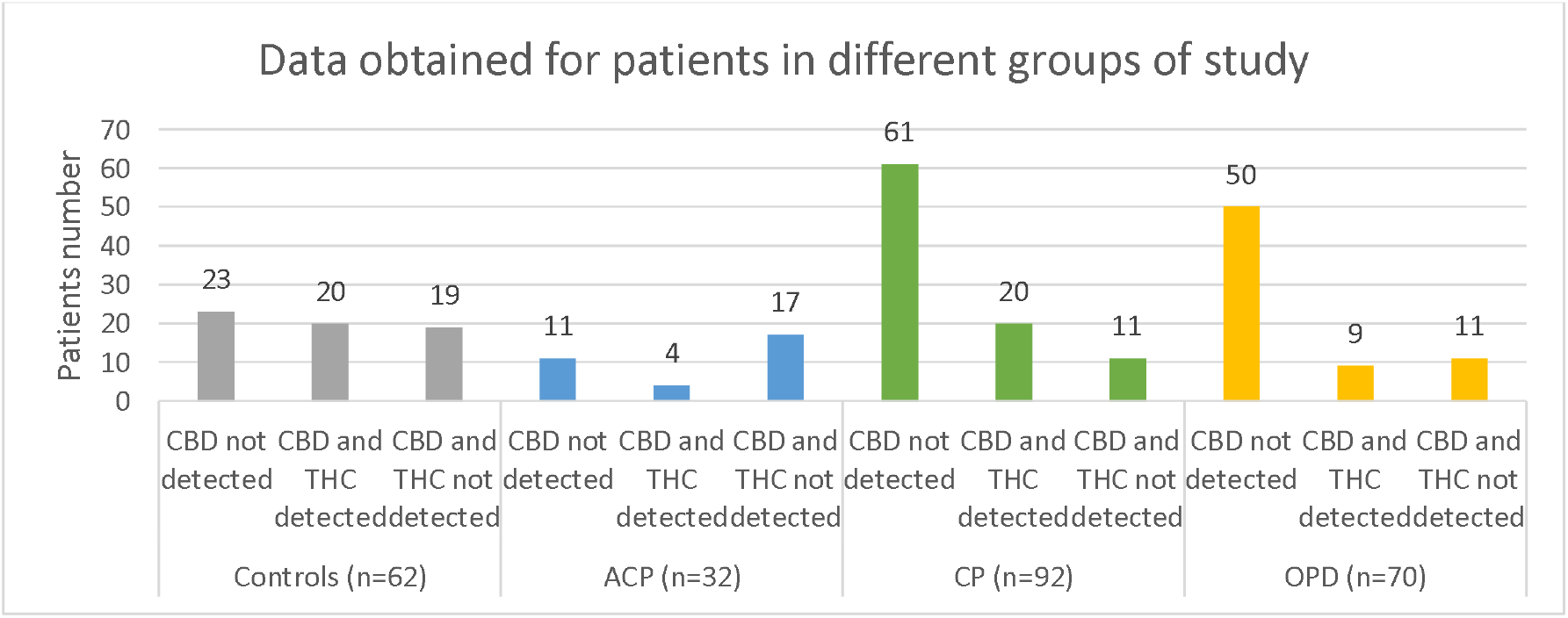
Distribution of data obtained concerning THC and CBD detection for patients of different groups. Undetected amounts were arbitrarily set at 0.0024 ng/mg. (Layout: Excel®)

The THC, CBD, and CBD/THC ratios between the different groups were compared using Tukey’s test (multiple comparisons). The stars in *Table 1* correspond to a significant difference between the two groups.

**Table 1.** Summary of comparisons made between different groups of patients. THC, CBD data and CBD/THC ratios are compared to each other.

|  | Control (n=62) |  |  | ACP (n=32) |  |  | CP (n=92) |  |  | OPD (n=70) |  |  |
| --- | --- | --- | --- | --- | --- | --- | --- | --- | --- | --- | --- | --- |
|  | THC | CBD | CBD/THC | THC | CBD | CBD/THC | THC | CBD | CBD/THC | THC | CBD | CBD/THC |
| Control |  |  |  |  |  |  | ** |  | **** |  |  | ** |
| ACP |  |  |  |  |  |  | ** | * | **** |  |  | *** |
| CP | ** |  | **** | ** | * | **** |  |  |  | *** |  |  |
| OPD |  |  | ** |  |  | *** | *** |  |  |  |  |  |

\*: P-value < 0.05
\*\*: P-value < 0.01
\*\*\*: P-value < 0.001
\*\*\*\*: P-value < 0.0001
ACP, acute cannabis psychosis; CP, chronic psychosis; OPD, other personality disorders; n, number of patients in the group

Descriptive statistics of the obtained data are presented and listed (*Table 2*) and are represented in the histograms (*figures 2 and 3*). These descriptive statistics show the repartition of CBD and THC values for patients in each group.

**Table 2.** Summary of descriptive statistics of data obtained for the different groups of patients.

|  |  | Control | ACP | CP | OPD |
| --- | --- | --- | --- | --- | --- |
| THC | min | 0.002400 | 0.002400 | 0.002400 | 0.002400 |
|  | max | 4.000 | 1.800 | 12.80 | 3.800 |
|  | average | 0.4373 | 0.1558 | 1.2523 | 0.2933 |
|  | CI (95 %) | 0.2259 - 0.6488 | 0.01598 - 0.2956 | 0.7773 - 1.727 | 0.1612 - 0.4253 |
| CBD | min | 0.002400 | 0.002400 | 0.002400 | 0.002400 |
|  | max | 1.900 | 0.4400 | 0.8700 | 0.1700 |
|  | average | 0.1019 | 0.0318 | 0.04883 | 0.01498 |
|  | CI (95 %) | 0.03343 - 0.1705 | (-) 0.002104 - 0.06568 | 0.02016 - 0.07751 | 0.005986 - 0.02398 |
| CBD / THC | min | 0.0007869 | 0.007500 | 0.0001875 | 0.001846 |
|  | max | 2.000 | 1.000 | 1.160 | 1.063 |
|  | average | 0.4874 | 0.5945 | 0.1910 | 0.2280 |
|  | CI (95 %) | 0.3623 - 0.6126 | 0.4303 - 0.7587 | 0.1195 - 0.2625 | 0.1351 - 0.3209 |

**Figure 2.**
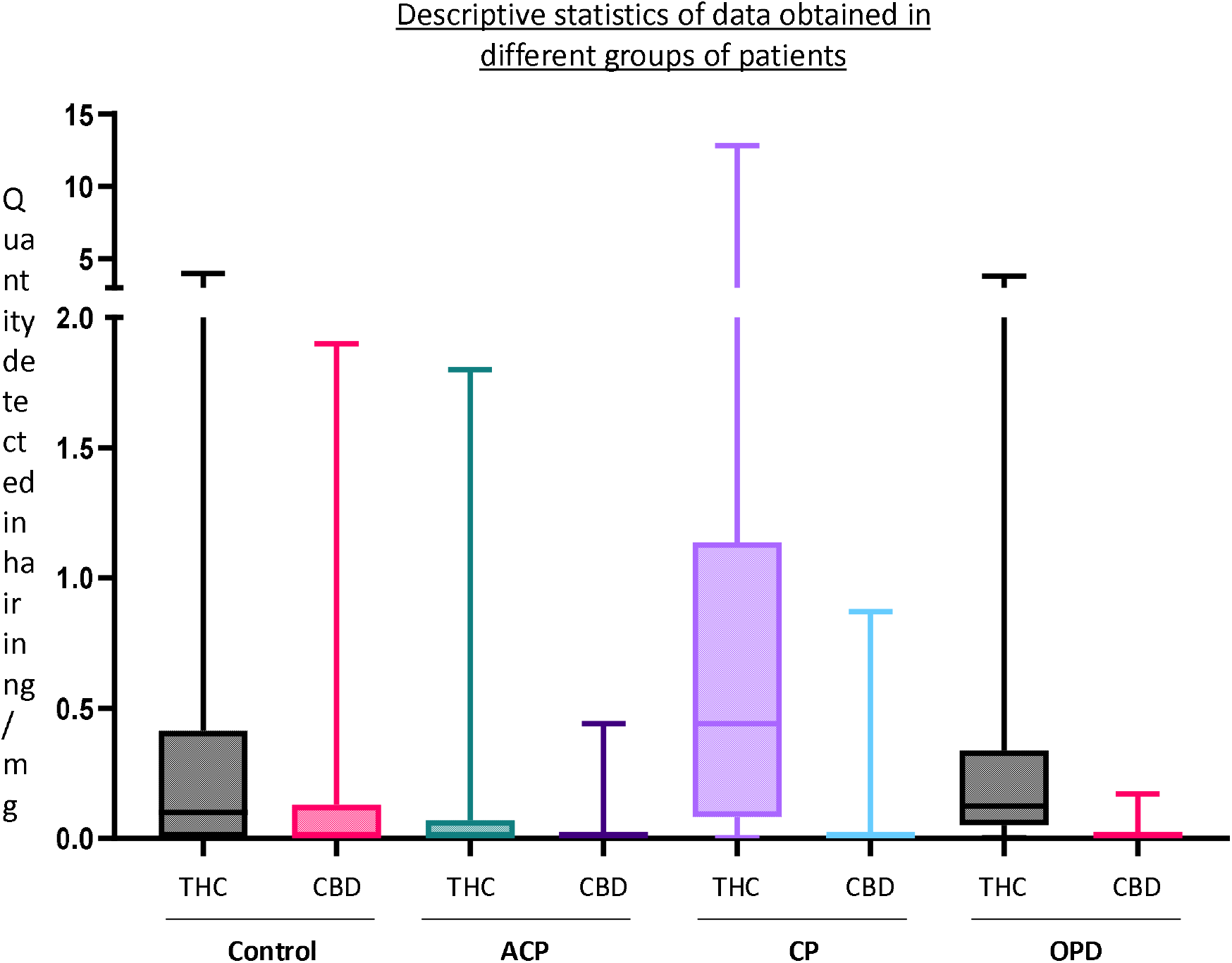
Descriptive statistics (obtained with GraphPad Prim 8®) of the concentrations of THC and CBD in ng/mg found in patients’ hair in different groups. Are represented on the boxplot: median and percentiles (25 and 75 %), minimum and maximum values (lower and upper limits, respectively, of the bar).

**Figure 3.**
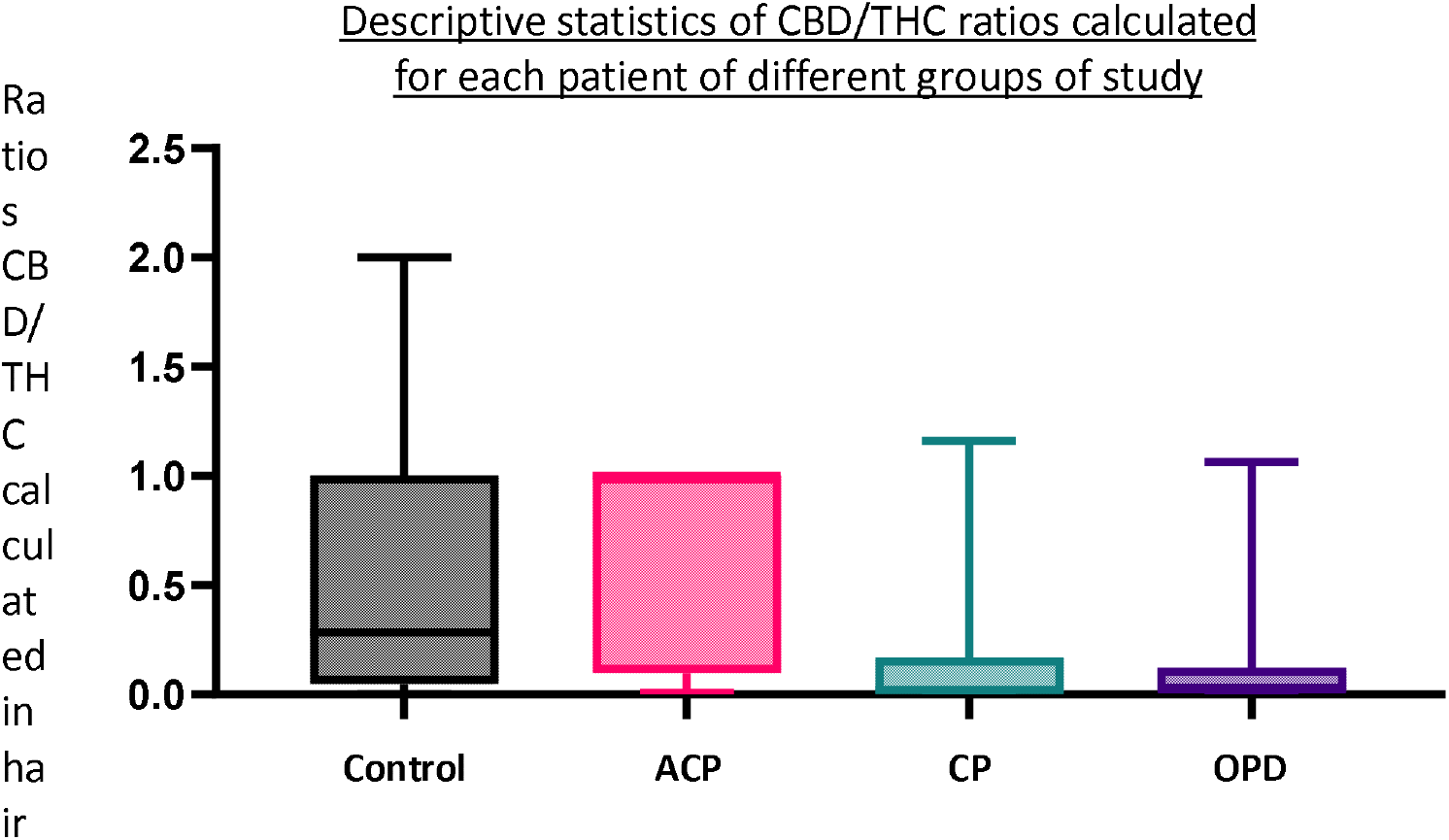
Descriptive statistics (obtained with GraphPad Prim 8®) of the CBD/THC ratios calculated for each patient in different groups. The boxplot shows median and percentiles (25 and 75 %), minimum and maximum values (lower and upper limits, respectively, of the bar).

## Discussion

From these analyses, the CP group was composed of people who possessed high levels of THC in their hair. This data may be correlated with the fact that patients with CP smoke more joints, or smoke more potent cannabis joints than people from other groups [17, 18]. Moreover, less CBD was found in the CP group than in the control group, and the CBD/THC ratio was significantly lower in the CP group than in the control group. Do patients in the CP group metabolize CBD in a particular way, or do these patients choose cannabis with a high psychotropic potency? The second hypothesis is the most likely [17, 18]. People who developed ACP are “light” consumers of cannabis and people from OPD group have approximately the same amount of THC found in hair than the control group. In this case, the quantity of CBD is less important in the OPD group than in the control group. CBD may play a role in the protection against chronic or periodic mental disease development [19, 20].

From these different data, it was possible to establish a decision-making tree (*figure 4*) that can guide clinicians in the early diagnosis of chronic psychosis in cannabis users admitted for BPD, and whose amounts of CBD and/or THC are either or not detectable or quantifiable in hair.

**Figure 4.**
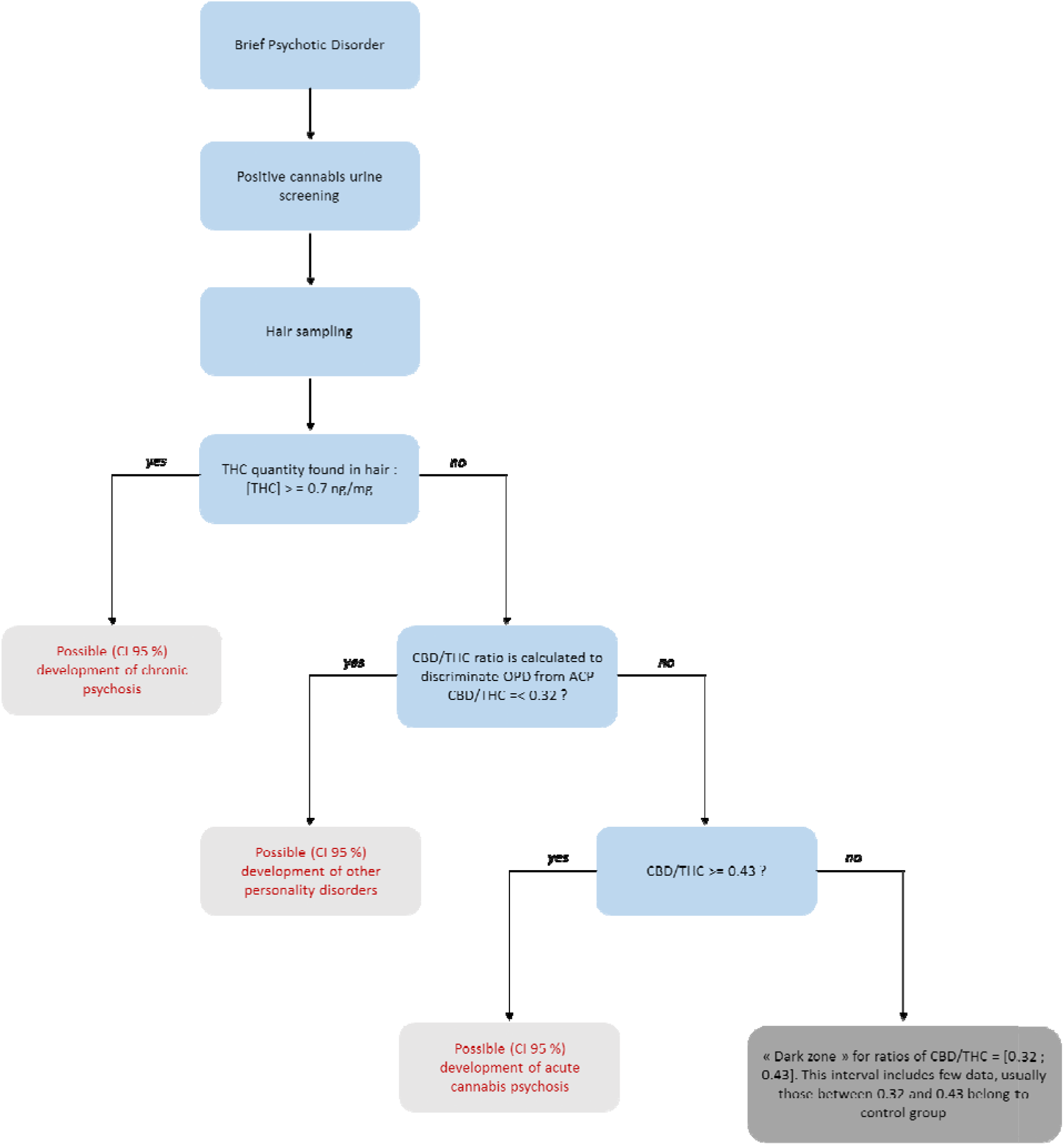
Decision-making tree constructed from the various statistical data obtained. ACP: acute cannabis psychosis; OPD: other personality disorders; CI: confidence interval

This study must continue to obtain larger patient and control groups; however, the trends are already emerging. In this study, a “high” level of THC detected (> 0.7 ng/mg) associated with a low CBD/THC ratio (< 0.26) are two parameters that taken together could be good markers of CP development. For OPD and ACP, discriminating between the two types of disease development is more ambiguous. Indeed, values for THC and CBD found in hair overlap and cannot discriminate between these two groups, even though THC seems to be lower and CBD higher in the ACP group than in the OPD group. It is necessary to calculate the CBD/THC ratios for patients to discriminate ACP from OPD. The ratios were higher in the ACP group (> 0.43) than in the OPD group (< 0.32).

This study highlights, once again, the protective role of CBD against the deleterious effects of THC [19, 20]. CBD is a cannabinoid with no or very low psychoactive effect, with which sedative (at high doses), anxiolytic, antiemetic, antiepileptic, antidystonic, and anti-inflammatory effects have been observed [21, 22]. This appears to antagonize the psychoactive effects of THC. Its mechanism of action is unclear, but it has been suggested that CBD may be an inverse agonist of cannabinoid receptors [21, 22], and that CBD shows an allosteric interaction with 5-HT_1A_ receptors [23].

## Conclusion

In association with clinical evaluation, this decision tree could be helpful for diagnosis and would allow early management of BPD in cannabis consumers. For a consumer who does not present with a psychiatric disorder, this decision tree would also make it possible to focus on his consumption to raise awareness. If necessary, the medical staff, patient’s entourage, and patient could be informed about the possibility of belonging to a group of patients at high risk of psychiatric decompensation. This provides an additional argument for efforts to control cannabis consumption by patients. This decision tree may provide more motivation to stop. Indeed, psychological education alone appears to be less effective in reducing cannabis consumption [24]. Such a decision tree, which may be adapted according to the regions of the world and consumer populations, would allow the patient to visualize the risk associated with heavy cannabis consumption. Such an approach could be included in behavioral treatment modalities for cannabis use disorders [24].

## Data Availability

All data produced in the present study are available upon reasonable request to the authors

## Acknowledgments

The authors are grateful to the volunteers who participated in the study. Students in the medical, pharmaceutical, and health sciences who participated in the collection of data and hair samples. For carrying out the capillary assays, the authors would like to thank ChemTox Laboratory, Illkirch-Graffenstaden (France) and more particularly, Dr Vincent Cirimele and Dr Olivier Roussel.

## Author Contributions

GS and J-YC contributed to the design, implementation of the research and made the clinical diagnosis, LC to the analysis of the results and to the writing of the manuscript. YB conceived the original, supervised the project and wrote the manuscript.

## Conflict of interests

The authors declare that they have no conflict of interest related to this article.

## Funding Statement

Not applicable.

## Ethical Compliance

All procedures were in accordance with the ethical standards of the Gaston Bourret Territorial Hospital research committee and with the 1964 Helsinki Declaration and its later amendments.

## Data Access Statement

Research data supporting this publication are available upon request.

## Notes

### Competing Interest Statement

The authors have declared no competing interest.

### Funding Statement

This study did not receive any funding

### Author Declarations

Ethic Committee of New Caledonia Territorial Center gave ethical approval for this work

### Summary of Updates

Updated abbreviations and clarified detection limits

